# Caring Under Pressure: A Qualitative Study of Nurse Job Satisfaction in a Private Hospital in Nigeria

**DOI:** 10.1101/2025.10.27.25338749

**Authors:** Mathew Ugochukwu Ugwuegbulam, Ejura Yetunde Salihu, Yamikani Nkhoma, Saima Tasneem

## Abstract

Using the Herzberg’s Two-Factor Theory, this study explored factors influencing job satisfaction among nurses in a private hospital in Nigeria. Ten nurses were recruited through purposive sampling to participate in interviews. Data was analyzed in MAXQDA using a thematic analysis approach. Extrinsic (hygiene) factors (i.e., high workload, low salary, limited opportunities for career advancement) were noted by respondents as significant sources of dissatisfaction, while intrinsic (motivational) factors (i.e., autonomy, peer support, and recognition) enhanced job satisfaction. Findings offer insights into how systemic and interpersonal factors influence nurse satisfaction and provide evidence-based strategies for enhancing retention in low-resource, profit-driven healthcare environments.

## Introduction

There is a rising demand for nurses and social care services worldwide due to the aging population and rising rates of chronic illnesses in all age gropus.^1^ The nursing workforce itself is also aging and experiencing high turnover globally.^1–4^ Compounding this issue is the migration of thousands of Nigerian nurses annually to countries such as the United Kingdom and the United States.^5–8^ High nurse turnover poses a serious challenge, especially in low-income countries that are experiencing already at the brink of crisis in healthcare sector due to limited resources. There is therefore a bigger demand for healthcare providers such as nurses, making job satisfaction a key concern in recruitment and retention.^5,9,10^

Job satisfaction and stress are key factors that determine nurse turnover rates. Job satisfaction includes not only emotional responses to work but also the extent to which job expectations are met.^10,11^ Research has established a strong relationship between worker satisfaction and the quality of patient care, as satisfied nurses tend to be more productive, creative, and committed.^12– 14^ So, from both patient and management perspectives, improving job satisfaction is essential for optimal work performance and patient outcomes.^15–17^

Herzberg’s Two-Factor theory states that there are two factors that drive job satisfaction and work performance. These factors are 1) hygiene (extrinsic) factors, which are related to the job itself, such as salary, policies, supervision, work environment, and job stability, and 2) motivators (intrinsic), such as recognition and personal fulfillment. The theory posits that both factors drive job satisfaction and retention.^18–20^

Although job satisfaction among nurses has been widely studied, critical gaps remain regarding job satisfaction, engagement, and retention among nurses, especially in the private sector in Nigeria.^21^ The World Health Organization highlights the need for more evidence on health worker retention strategies in low-income settings to support progress toward the Sustainable Development Goals.^22^

This study expands on the existing body of research by identifying factors associated with satisfaction or dissatisfaction in the Nigerian healthcare workplace context, using Herzberg’s two-factor theory as a guiding framework.

## Methods

Using purposive sampling, ten registered nurses were recruited from five departments (Maternity/Labor and Delivery Ward, Surgical Ward, Emergency Department, Intensive Care Unit, and Pediatric Ward) of a private hospital in Southwestern Nigeria to participate in one-on-one semi-structured interviews. Interviews were led by an experienced qualitative researcher and held in-person in a private room.^23^ Each session lasted for 45-60 minutes. See Appendix A for the interview guide.^24^

Interviews were audio-recorded and transcribed verbatim. The researchers analyzed the data using MAXQDA (VERBI Software, 2024).^25^ We conducted the analysis using Braun and Clarke’s six-phase framework for thematic analysis, which includes familiarization, generation of initial codes, theme searching, theme reviewing, theme defining, and report production.^26^

## Results

This section presents the sociodemographic profile of the participants who took part in the study (see Table 1).

**Table 1:** Gender Distribution of The Respondents.

| Gender | Frequency | Percentage % |
| --- | --- | --- |
| Male | 3 | 30 |
| Female | 7 | 70 |
| Total | 10 | 100 |

**Table 1** shows the gender distribution of the respondents. Of the ten participants, seven were female and three were male, reflecting the general gender composition commonly observed within the nursing profession in Nigeria.

Table 2 presents the age distribution of participants. One respondent (10%) was between the ages of 21 and 25. Two respondents (20%) each fell within the age ranges of 26 to 30 and 31 to 35. Three participants (30%) were aged between 36 and 40, while two respondents (20%) were over 40.

**Table 2:**
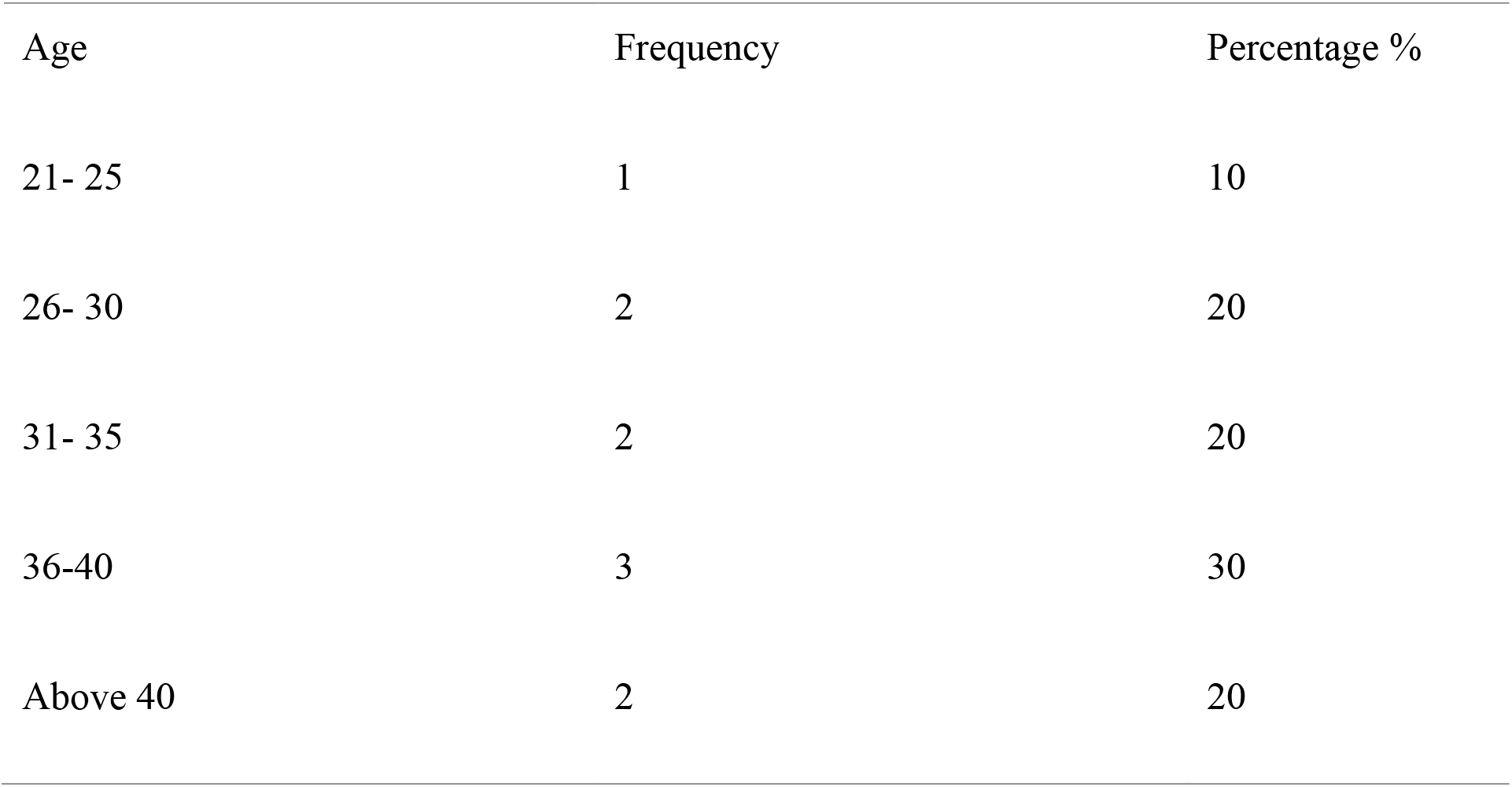
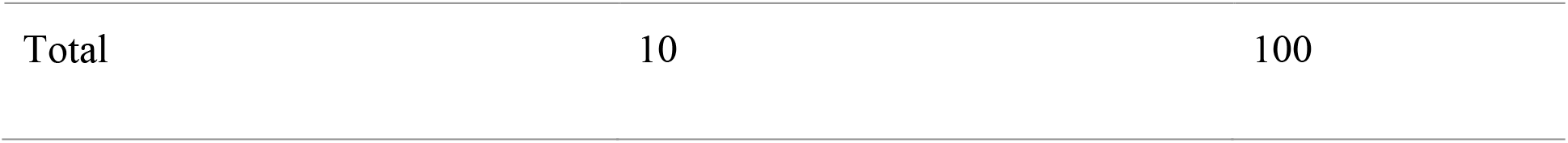
Age of Respondents.

| Age | Frequency | Percentage % |
| --- | --- | --- |
| 21- 25 | 1 | 10 |
| 26- 30 | 2 | 20 |
| 31- 35 | 2 | 20 |
| 36-40 | 3 | 30 |
| Above 40 | 2 | 20 |
| Total | 10 | 100 |

Table 3 below shows the distribution of respondents by years of professional experience. Three participants (30%) had between 1 and 5 years of experience, while two respondents (20%) reported between 6 and 10 years of experience. The majority of participants, five (50%), had more than 10 years of experience in the nursing profession.

**Table 3:** Respondents’ Years of Experience.

| Years of experience | Frequency | Percentage % |
| --- | --- | --- |
| 1-5 | 3 | 30 |
| 6-10 | 2 | 20 |
| Above 10 | 5 | 50 |
| Total | 10 | 100 |

## Qualitative Results

This section presents the themes from interviews. Each theme is supported by illustrative quotes from participants, providing insight into the lived experiences of nurses in the private health sector.

### Hygiene (Extrinsic) Factors: Barriers to Job Satisfaction

Theme 1: Perceived Inadequacy of Compensation. A recurring and strongly expressed concern among all participants was dissatisfaction with their salary.

#### One participant stated

“Honestly, the remuneration is poor. We do most of the jobs, and we are not given adequate pay. It is unfair to us, the nurses in this private sector.”

{Nurse 01, female}

#### Another added

“The salary is not encouraging compared to the workload and even that of other health workers. We will really love the managerial staff to do something about it.”

{Nurse 04, female}

Theme 2: Limited Opportunities for Career Growth. Opportunities for advancement and promotion emerged as a central theme in the interviews, mentioned by all respondents as a key factor influencing their job satisfaction.

#### One participant stated

“If the organization can train us and help us advance via certification programs, attending seminars, and training, I will be so glad, and it will help me give my best.”

{Nurse 10, male}

#### Another participant said

“I wish the management could do more by helping us advance our careers. Most of us have been at the same level for years since we came here. I have been for over 4 years, and I have not advanced. I do not mind leaving to get some professional certificates, but it is very expensive.”

{Nurse 9, female}

Theme 3: Excessive Workload and Burnout. A major contributor to dissatisfaction among participants was the excessive workload burden. Nurses reported high patient-to-staff ratios and described their shifts as physically and mentally draining.

“Most of us usually work a few hours in addition to our shift hours because the demand is just too much for us.”

{Nurse 09, female}

It is always a hectic routine for us because we are usually outnumbered by the patients ‘demands.

{Nurse 03, male}

### Motivators (Intrinsic): Contributors to Job Satisfaction

Theme 1: Supportive Peer Relationships as a Buffer. Supportive relationships among coworkers were highlighted as a source of job satisfaction. While peer support is extrinsic by nature, participants described how positive colleague relationships enhanced their sense of belonging and confidence, factors that align with Herzberg’s intrinsic motivators.

‘The difference between here and where I came from is that there is a good relationship among the nurses and other medical practitioners. One time I had a patient in a coma, being on a night shift, my colleagues came through and helped me with ideas which made me arrest the situation.”

{Nurse 1, Female}

“One thing that has helped me settle quickly here is my relationship with my coworkers. They helped me overcome my fear, and my weaknesses were not exposed. It helped me feel at home to do better.”

{Nurse 07, female}

Theme 2: Recognition as a Driver of Professional Fulfillment. Participants consistently expressed that recognition and appreciation from management were critical to their sense of fulfillment and professional identity.

#### One participant shared

“I remembered two years ago when I was given two awards at the end of the year for being the most punctual and most hardworking nurse. It made me feel so proud of myself and my job, and it also challenged my colleagues to be better.”

{Nurse 04, female}.

Theme 3: Autonomy and Decision-Making at Work. Autonomy emerged as a strong theme linked to job satisfaction. Participants emphasized that autonomy fosters self-esteem and competence, but they also shared that exercising independent judgment was sometimes met with disapproval from supervisors.

“I tend to perform better when I am allowed to make decisions regarding my tasks and act independently. It builds my self-esteem and increases satisfaction.”

{Nurse 08, female.}

“I hate it when my superiors get angry with me for making critical decisions on the job. I remembered that one day I had to sedate a patient before calling the doctor; even though it salvaged the situation, I was still given a query.”

{Nurse 10, male}

## Discussion

Findings revealed that the absence of key hygiene factors and the neglect of motivators contribute to diminished wellbeing and morale among nurses in private hospital settings.

Most of the respondents in this study noted that lack of institutional investment in the nursing workforce was a prominent concern. Respondents complained that majority of the opportunities for advancement (e.g., workshops or study leave to pursue postgraduate training) were reserved for only senior medical personnel. This lack of equity in professional development fosters resentment and contributes to low morale. Hospital administrators may find it helpful to introduce structured career ladder programs and more clearly defined roles, as these could bring clarity, increase motivation, and support better performance.

Another key concern was the excessive workload. Nurses reported being overburdened due to staff shortages, which led to burnout and extended work hours. Adding to the excessive workload is frustration, which they said stems from being given several responsibilities outside the scope of their job description. While some had voiced management concerns, they noted that little action was taken to address the issue. This result validates prior studies that show that high psychological distress is associated with high workload and inadequate staffing.^3,11,28^

Frustration about unclear job expectations has also been noted in prior studies in this population.^29–31^

The challenges noted by the participants in this study are validated by previous studies of healthcare workforce wellbeing in different workplace settings. Within the context of private hospitals in Nigeria, several systemic challenges were identified that have persisted over the years. While patients usually prefer private hospitals due to quicker access and shorter wait times,^21,32–34^ these operational advantages often come at the cost of nurse wellbeing. A possible explanation for this is that many private institutions are profit-driven so that stakeholders may limit investments in staff wellbeing, including low salaries, high nurse-to-patient ratios, limited staffing flexibility, limited benefits, and limited career development opportunities. In other healthcare settings, a similar trend has been observed.^28,35,36^ These studies found that environments characterized by long working hours, limited resources, and high direct care responsibilities are associated with higher stress levels and lower job satisfaction compared to better-resourced workplace settings. These findings highlight how workplace setting can amplify work-related stress and underscores the need for responsive policies that address both workload and structural inequities in private healthcare settings.

Several motivating factors were also identified. Autonomy, for instance, was mentioned as a strong contributor to job satisfaction, particularly for nurses who had more years of experience. All the respondents reported feeling more empowered when allowed to make independent clinical decisions. Many participants said they felt empowered when they were trusted to make independent clinical decisions. At the same time, several recalled being reprimanded by supervisors for taking such initiative, pointing to a gap between what was expected of them and the level of support they received from management. This reinforces the need for consistent policies on decision-making authority.

Similarly, positive relationships with coworkers also played a significant role in enhancing job satisfaction. Nurses described collegial support, emotional safety, and teamwork as key elements that helped them cope with stress and perform better. These findings align with previous studies,^37–39^ which emphasize the importance of interprofessional collaboration and peer support in improving workplace satisfaction.

Another central theme was recognition and appreciation. Respondents expressed their desire for acknowledgment and positive reinforcement from management. They mentioned that absence of recognition contributed to them feeling invisible, which led to disengagement, a key indicator of high turnover.^40,41^

Overall, study findings show that job satisfaction among nurses in Nigeria is multifactorial, shaped by a combination of structural, interpersonal, and psychological factors. A participatory management style, coupled with transparent performance appraisals and career development structures, could significantly improve retention and performance in private hospitals in Nigeria.

## Limitations

The sample size (10 registered nurses from a single private hospital) is small. Therefore the result from this study is not generalizable to other hospital settings in Nigeria. Although participants were drawn from different departments, their experiences may not reflect the broader spectrum of nurses working in other private or public hospitals. Interviews were conducted in one session. Having multiple sessions spread over a period of time might have generated insights that show evolving perceptions over time. Despite these limitations, the study offers critical preliminary insights that helps us better understand the factors influencing job satisfaction among nurses in Nigeria’s private health sector.

## Conclusion

High migration pattern among Nigerian-trained nurses has left the local healthcare system particularly vulnerable, underscoring the need for sustainable staff retention strategies. To retain talented nurses, hospital administrators should ensure that the workplace is supportive, and policies are designed to allow nurses’ autonomy in clinical decision-making. Administrators could also offer structured mechanisms for compensating nurses. Addressing these issues through participatory management, equitable investment in staff professional development, and fair workload distribution will enhance nurse retention and performance, ultimately improving the quality of patient care in Nigeria’s private health sector.

## Data Availability

All data produced in the present study are available upon reasonable request to the authors

## Appendix A: Interview Questions

1. Can you tell me about your experience working as a nurse in a private hospital?
  a. What has been most rewarding or most challenging about this experience?
2. What expectations did you have before becoming a registered nurse?
  a. How did you imagine the role before you started?
  b. In what ways have your actual experiences matched or differed from those expectations?
3. How would you describe your overall feelings about your current job?
  a. What aspects of your work do you enjoy the most, and what aspects do you find challenging?
4. How long do you see yourself continuing to work in this hospital?
  a. What factors make you want to stay longer?
  b. What might make you consider leaving sooner?

